# Association Between Preoperative Albumin Corrected Anion Gap and Postoperative Delirium in Cardiac Surgery Patients

**DOI:** 10.64898/2026.05.07.26352646

**Authors:** Mostafa Abbas, Thomas Morland, Rohit Sharma, Neria Bitton, Maya Lichtenstein, H. Lester Kirchner, Scott LeMaire, Yasser El-Manzalawy

## Abstract

**Background:** Delirium, a common and multifactorial complication after cardiac surgery, is influenced by several factors including inflammation, metabolic disturbances, and cerebral hypoperfusion. Because these factors can be reflected in an elevated anion gap (AG), we hypothesized that a higher preoperative albumin corrected anion gap (ACAG) is associated with increased risk of delirium and 1-year mortality after cardiac surgery.

**Methods:** We examined a retrospective cohort of adult patients within our healthcare system who underwent cardiac surgery between 2014 and 2022 and had a recorded Confusion Assessment Method for the ICU (CAM-ICU) evaluation. Patients were excluded if they had documented preoperative delirium during the index hospital admission or a history of dementia. The final cohort included 4,482 patients. Preoperative laboratory values were collected, using the most recent results obtained within 48 hours prior to surgery. The primary outcome was delirium after cardiac surgery (DACS), defined as delirium occurring within postoperative days 1 through 5. The secondary outcome was all-cause 1-year mortality.

**Results:** The incidence of DACS and 1-year mortality were 9.5% and 4.8%, respectively. A multivariable logistic regression model adjusting for baseline characteristics showed that higher ACAG was significantly associated with higher risk of DACS (adjusted odds ratio (AOR) = 1.56, 95% Confidence Interval (CI) = 1.40-1.74, p < 0.001). Other predictors of DACS included increasing age (AOR = 1.31, CI = 1.16-1.48, p < 0.001), surgery duration (AOR = 1.35, CI = 1.22-1.49, p < 0.001), and history of delirium (AOR = 1.70, CI = 1.29-2.24, p < 0.001). Moreover, increasing ACAG was also associated with 1-year mortality (AOR = 1.35, CI = 1.16-1.56, p < 0.001). Finally, receiver operating characteristic (ROC) analysis demonstrated that ACAG exhibited superior predictive performance compared with AG and anion gap to bicarbonate ratio (AGBR) for both DACS and 1-year mortality outcomes.

**Conclusions:** Higher preoperative ACAG was associated with elevated risk for DACS and 1-year mortality. Preoperative ACAG is an accessible and cost-efficient biomarker that may improve risk stratification for cardiac surgery patients.

## Introduction

Delirium after cardiac surgery (DACS) is a frequent complication, with incidence rates ranging from 10% to 21% [1-7]. Patients undergoing cardiac surgery are at higher risk of postoperative delirium compared to patients undergoing non-cardiac surgery due to a higher prevalence of risk factors including older age, cerebrovascular disease, diabetes, renal disease, and intraoperative cardiac surgery specific risk factors such as cardiopulmonary bypass [1, 7, 8]. DACS has been associated with higher rates of mortality [9, 10], increased duration of hospitalization [10, 11], prolonged duration of mechanical ventilation [12, 13], and long-term cognitive impairment [14, 15]. Despite its high prevalence and serious consequences, the underlying pathophysiologic mechanisms of DACS remain poorly understood [16]. Current approaches to early identification and prevention of DACS are limited, which is an urgent concern given that up to 40% of cases may be preventable with timely and targeted interventions [17-20].

Preoperative or initial ICU admission laboratory measurements offer a simple, easily available, and inexpensive source of biomarkers for risk stratification of postoperative and ICU-related outcomes [21-23]. For example, the serum anion gap (AG), defined as the difference between unmeasured anions and unmeasured cations, is an easily calculated fundamental indicator of a patient’s metabolic status [24, 25]. Recently, AG has emerged as a valuable prognostic marker of mortality across various critical care settings, including patients with acute kidney injury (AKI) [21, 26], acute myocardial infarction (AMI) [22], and congestive heart failure (CHF) [21, 23, 27]. However, AG levels are strongly influenced by albumin, a major unmeasured anion [28]. Hypoalbuminemia can lead to underestimation of AG levels. To address this limitation, several corrected or composite AG metrics have been developed to provide more accurate and reliable estimates of the metabolic state, such as albumin corrected anion gap (ACAG)[23, 28, 29], anion gap to calcium ratio (AGCR) [30], and anion gap to bicarbonate ratio (AGBR) [31].

Because DACS may result from multiple factors (including inflammation, metabolic disturbances, and cerebral hypoperfusion) often reflected by an elevated AG, we hypothesized that a higher preoperative ACAG is associated with an increased risk of postoperative delirium and 1-year mortality following cardiac surgery.

## Methods

### Study population

We conducted a retrospective cohort study within Geisinger health system, a large integrated healthcare network in Pennsylvania, USA. The study protocol was approved by the Geisinger Institutional Review Board (Approval No: 2020–0101) with a waiver of informed consent. We identified adults who underwent coronary artery bypass grafting (CABG), aortic valve replacement, or mitral valve replacement between 2014 and 2022, and who had a Confusion Assessment Method for the ICU (CAM-ICU) assessment documented during their postoperative course. After excluding patients with preoperative delirium at index admission or with a history of dementia, our final cohort comprised 4,482 patients.

### Clinical variables and outcomes

Patients’ electronic health records (EHR) were linked with our institutional Society of Thoracic Surgery (STS) database in order to enrich our analysis with additional clinical variables. Patient comorbidities were determined using the Charlson Comorbidity Index based on the International Classification of Diseases (ICD) diagnosis codes. History of atrial fibrillation (AF) was defined as a documented AF in patient records including ECG findings and diagnosis codes. Preoperative laboratory values were defined as the most recent measurements obtained within 48 hours prior to surgery. To account for underestimated levels of preoperative serum AG due to hypoalbuminemia, which is common among cardiac surgery patients, ACAG was computed as follows [28, 32]: *ACAG* (*mmol*⁄*L*) = *AG* + [44 − *albumin*(*g*⁄*L*)] *x* 0.25. The primary outcome was DACS, defined as a documented delirium occurring within postoperative days 1 through 5. Delirium was defined as a positive CAM-ICU assessment in patients with a Richmond Agitation-Sedation Scale (RASS) score ≥ –3. The secondary outcome was postoperative all-cause 1-year mortality.

### Statistical analysis

Categorical variables were reported as percentages, and the chi-square test was used to assess whether two (or more) proportions were different from each other. Missing values for continuous variables were imputed using the median of observed values. Imputed continuous variables were then summarized as medians and interquartile ranges (IQR). Multivariable logistic regression was used to evaluate the associations between baseline variables and the outcomes of interest. Before model fitting, all continuous variables were standardized using z-transformation. Two multivariable models were specified. Model^a^ included ACAG as a continuous variable, whereas Model^b^ modeled ACAG categorically using three predefined groups (low, medium, and high) based on the 1^st^ and 3^rd^ quartiles of the ACAG measurements in Table 1. Both models were adjusted for the same set of covariates listed in Table 1. To account for multiple testing across baseline variables, Bonferroni-adjusted p-values (q-values) were reported. Survival analysis using Kaplan-Meier (KM) and Cox Proportional Hazard model was applied to characterize survivals patterns in DACS and non-DACS groups. Differences between KM curves were assessed using the log-rank test. All analyses were conducted using R version 4.4.2 (R Foundation for Statistical Computing, Vienna, Austria), and a two-sided p-value <0.05 was considered statistically significant.

**Table 1:** Baseline characteristics of the study cohort stratified by ACAG categorized into low, medium and high based on the 1^st^ and 3^rd^ quartiles.

| **Variable** | **Low ACAG**<br>**(n = 1122)** | **Medium ACAG**<br>**(n = 2235)** | **High ACAG**<br>**(n = 1125)** | **Total**<br>**(n = 4482)** | **q-value** |
| --- | --- | --- | --- | --- | --- |
| Age (years) | 65 (58, 72) | 67 (59, 73) | 67 (59, 74) | 66 (59, 73) | 0.002 |
| Female | 226 (20.1%) | 704 (31.5%) | 347 (30.8%) | 1277 (28.5%) | <0.001 |
| BMI (Kg/m<sup>2</sup>) | 30.1 (26.7, 34.7) | 30 (26.3, 34.6) | 29.7 (26, 34) | 30 (26.3, 34.5) | 1.000 |
| Emergency admission | 167 (14.9%) | 361 (16.2%) | 450 (40%) | 978 (21.8%) | <0.001 |
| CABG and valve | 93 (8.3%) | 239 (10.7%) | 92 (8.2%) | 424 (9.5%) | 0.494 |
| Surgery duration (hours) | 3.6 (2.9, 4.6) | 3.9 (3.1, 4.8) | 4 (3.2, 5) | 3.8 (3.1, 4.8) | <0.001 |
| Ejection fraction (%) | 57 (50, 60) | 57 (47, 61) | 52 (42, 57) | 57 (47, 60) | <0.001 |
| Congestive heart failure | 373 (33.2%) | 873 (39.1%) | 521 (46.3%) | 1767 (39.4%) | <0.001 |
| Myocardial infarction | 401 (35.7%) | 852 (38.1%) | 658 (58.5%) | 1911 (42.6%) | <0.001 |
| PVD | 347 (30.9%) | 720 (32.2%) | 352 (31.3%) | 1419 (31.7%) | 1.000 |
| COPD | 501 (44.7%) | 1062 (47.5%) | 530 (47.1%) | 2093 (46.7%) | 1.000 |
| Renal disease | 174 (15.5%) | 488 (21.8%) | 323 (28.7%) | 985 (22%) | <0.001 |
| Diabetes | 404 (36%) | 1016 (45.5%) | 598 (53.2%) | 2018 (45%) | <0.001 |
| Cerebrovascular disease | 226 (20.1%) | 509 (22.8%) | 279 (24.8%) | 1014 (22.6%) | 0.778 |
| Hypertension | 968 (86.3%) | 2000 (89.5%) | 1038 (92.3%) | 4006 (89.4%) | <0.001 |
| History of AF | 297 (26.5%) | 593 (26.5%) | 394 (35%) | 1284 (28.6%) | <0.001 |
| History of delirium | 117 (10.4%) | 261 (11.7%) | 159 (14.1%) | 537 (12%) | 0.552 |
| Bilirubin (mg/dL) | 0.5 (0.4, 0.7) | 0.5 (0.4, 0.7) | 0.5 (0.4, 0.7) | 0.5 (0.4, 0.7) | 1.000 |
| Creatinine (mg/dL) | 1 (0.8, 1.1) | 1 (0.8, 1.2) | 1 (0.8, 1.2) | 1 (0.8, 1.2) | 1.000 |
| Glucose (mg/dL) | 114 (103, 133) | 117 (106, 137) | 122 (104, 152) | 117 (104, 139) | <0.001 |
| Hematocrit (%) | 38.1 (35, 41.3) | 37.4 (33.6, 40.9) | 37 (32, 40.5) | 37.5 (33.9, 41) | <0.001 |
| Hemoglobin (g/dL) | 14 (13, 14.9) | 13.5 (12.4, 14.5) | 13 (11.5, 14.3) | 13.5 (12.3, 14.6) | <0.001 |
| Prothrombin time-INR | 1 (1, 1.1) | 1 (1, 1.1) | 1.1 (1, 1.1) | 1 (1, 1.1) | <0.001 |
| HbA1C (%) | 5.8 (5.4, 6.4) | 5.9 (5.5, 6.8) | 6 (5.6, 7.2) | 5.9 (5.5, 6.8) | <0.001 |
| PWR | 27.1 (21.7, 32.7) | 27.6 (22.3, 34.1) | 25.9 (20.3, 32.1) | 27.1 (21.7, 33.3) | <0.001 |
| ACAG (mmol/L) | 6.2 (4.9, 7) | 9.1 (8.3, 9.8) | 13 (12, 14.5) | 9.1 (7.6, 11.2) | <0.001 |
| **Outcomes** |  |  |  |  |  |
| DACS | 48 (4.3%) | 196 (8.8%) | 184 (16.4%) | 428 (9.5%) | <0.001 |
| 1-year mortality | 27 (2.4%) | 91 (4.1%) | 97 (8.6%) | 215 (4.8%) | <0.001 |

## Results

### Baseline characteristics

A total of 4,482 adult cardiac surgery patients were included in our retrospective study cohort, of whom 1277 (28.5%) were females. Because 98% of these patients were White, we did not include race or ethnicity in the analysis. The incidence of DACS and 1-year mortality were 9.5% and 4.8%, respectively. **Table 1** summarizes the baseline characteristics of the study cohort stratified by ACAG categorized into low, medium and high based on the 1^st^ and 3^rd^ quartiles. The prevalence of DACS and 1-year mortality were approximately doubled in the high ACAG group compared with low and medium ACAG groups. Patients in the high ACAG group had longer surgery durations and had a greater prevalence of comorbidities.

### Association between ACAG and DACS

Table 2 summarizes the significant predictors of DACS identified using two multivariable logistic regression models, Model^a^ and Model^b^, both adjusted for all covariates listed in Table 1. The only difference between the two models is how ACAG was modeled. In Model^a^, ACAG was modeled as a continuous variable, while in Model^b^ ACAG was grouped into three categories, low medium, and high. Both models identified the same set of significant predictors. In both analyses, elevated ACAG levels were associated with an increased risk of DACS. In addition to ACAG, INR was the only other preoperative laboratory variable that independently predicted DACS.

**Table 2:** Significant predictors of DACS.

| **Model<sup>a</sup>** |  |  | **Model<sup>b</sup>** |  |  |
| --- | --- | --- | --- | --- | --- |
| **Variable** | **OR (95% CI)** | **p-value** | **Variable** | **OR (95% CI)** | **p-value** |
| Age | 1.31 (1.16-1.48) | <0.001 | Age | 1.30 (1.15-1.47) | <0.001 |
| CABG and valve | 1.40 (1.03-1.91) | 0.034 | CABG and valve | 1.39 (1.02-1.9) | 0.036 |
| Surgery duration | 1.35 (1.22-1.49) | <0.001 | Surgery duration | 1.35 (1.22-1.49) | <0.001 |
| PVD | 1.32 (1.05-1.65) | 0.016 | PVD | 1.31 (1.05-1.64) | 0.016 |
| History of delirium | 1.70 (1.29-2.24) | <0.001 | History of delirium | 1.69 (1.28-2.23) | <0.001 |
| INR | 1.08 (1.01-1.16) | 0.025 | INR | 1.10 (1.02-1.18) | 0.013 |
| ACAG | 1.56 (1.40-1.74) | <0.001 | Medium ACAG | 2.00 (1.43-2.79) | <0.001 |
|  |  |  | High ACAG | 3.83 (2.69-5.43) | <0.001 |

Model<sup>a</sup> and Model<sup>b</sup> refer to multivariable regression analyses with ACAG modeled as a continuous and a discretized variable, respectively.

### Association between ACAG and 1-year mortality

Table 3 reports significant predictors of postoperative 1-year mortality obtained using Model^a^ and Model^b^. Both models demonstrated association between higher preoperative ACAG and elevated risk of 1-year mortality. Consistently, the two models showed that increased risk of 1-year mortality is associated with older age, female sex, prolonged surgery duration, lower ejection fraction, congestive heart failure, history of AF, lower preoperative hemoglobin, and higher preoperative bilirubin and INR levels. **Figure 1** illustrates the 1-year survival patterns for patients in the low, medium, and high ACAG groups.

**Table 3:**
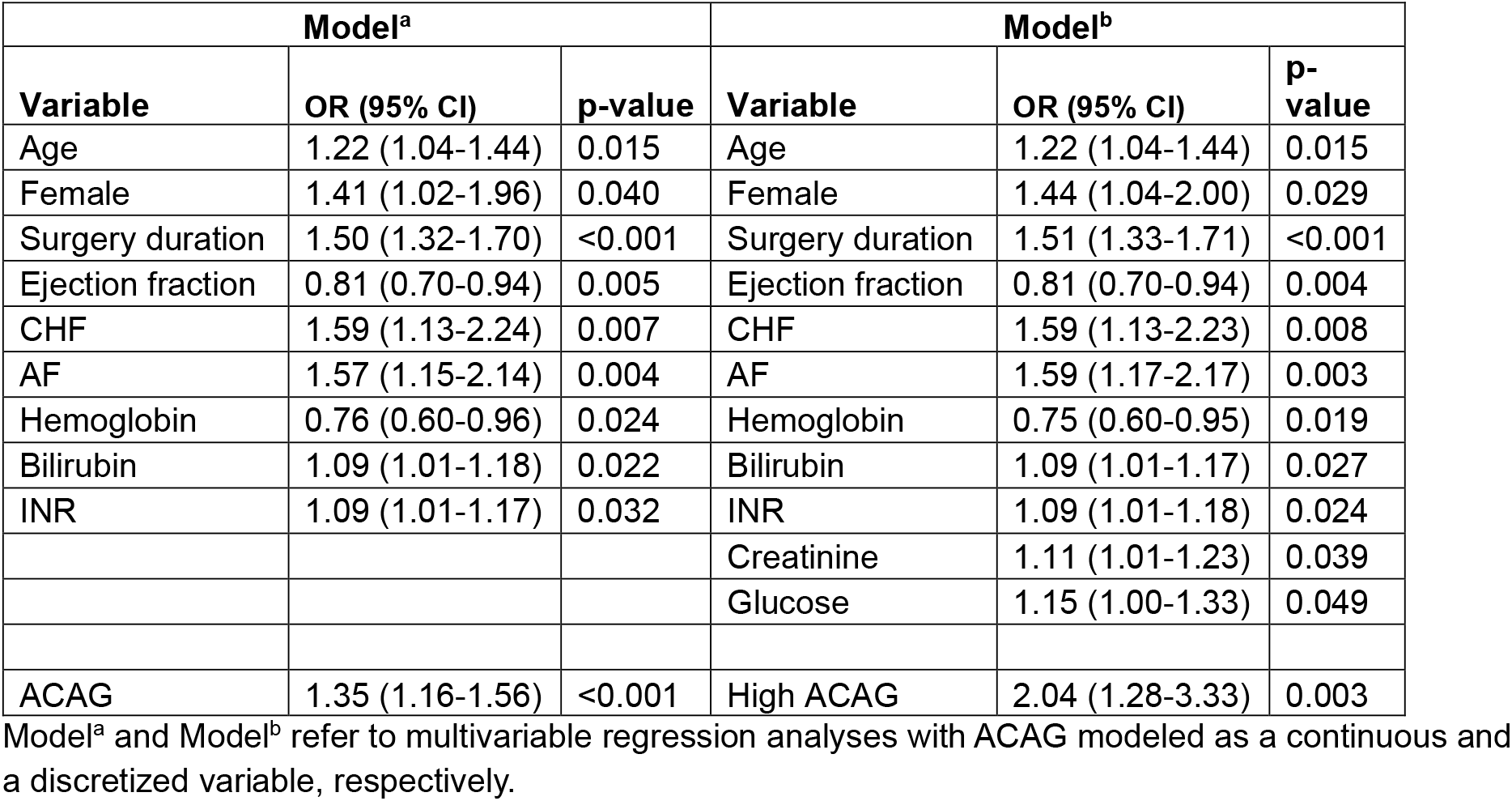
Significant predictors of postoperative 1-year mortality.

**Figure 1:**
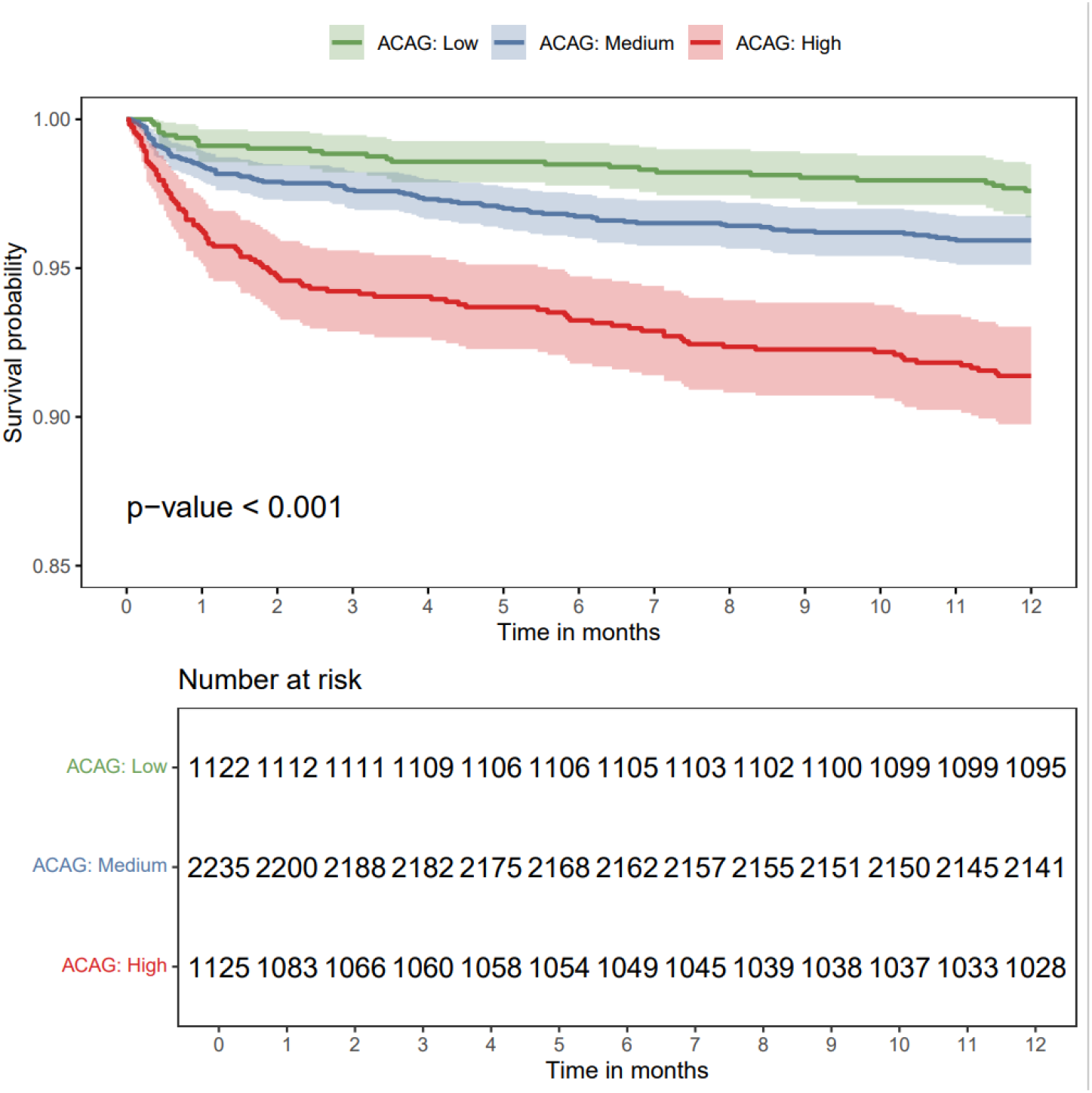
Kaplan–Meier survival curves over 1 year with 95% confidence intervals for patients stratified by their preoperative ACAG group.

### Sensitivity analysis

For sensitivity analyses, two additional multivariable logistic regression models were fitted using the Model^a^-based analysis, in which ACAG was replaced with measured preoperative AG and AGBR, respectively. Table 4 compares the results of these Model^a^-based analyses for ACAG, AG, and AGBR in assessing their associations with DACS and 1-year mortality. Overall, all three AG measurements were independently associated with an elevated risk of DACS and 1-year mortality, with ACAG demonstrating the strongest association. Figure 2 compares the receiver operating characteristic (ROC) curves of the three AG-based measurements for predicting DACS and 1-year mortality. For both outcomes, ACAG consistently demonstrated superior predictive performance with an AUC score of 0.65.

**Table 4:** Associations of preoperative ACAG, AG, and AGBR continuous variables with DACS and 1-year mortality based on multivariable logistic regression models adjusted for the covariates in Table 1.

| Variable | OR (95% CI) | p-value | OR (95% CI) | p-value |
| --- | --- | --- | --- | --- |
| DACs |  |  | 1-year mortality |  |
| ACAG | 1.56 (1.40-1.74) | <0.001 | 1.35 (1.16-1.56) | <0.001 |
| AG | 1.45 (1.31-1.61) | <0.001 | 1.23 (1.07-1.42) | 0.004 |
| AGBR | 1.32 (1.19-1.45) | <0.001 | 1.20 (1.05-1.37) | 0.009 |

**Figure 2:**
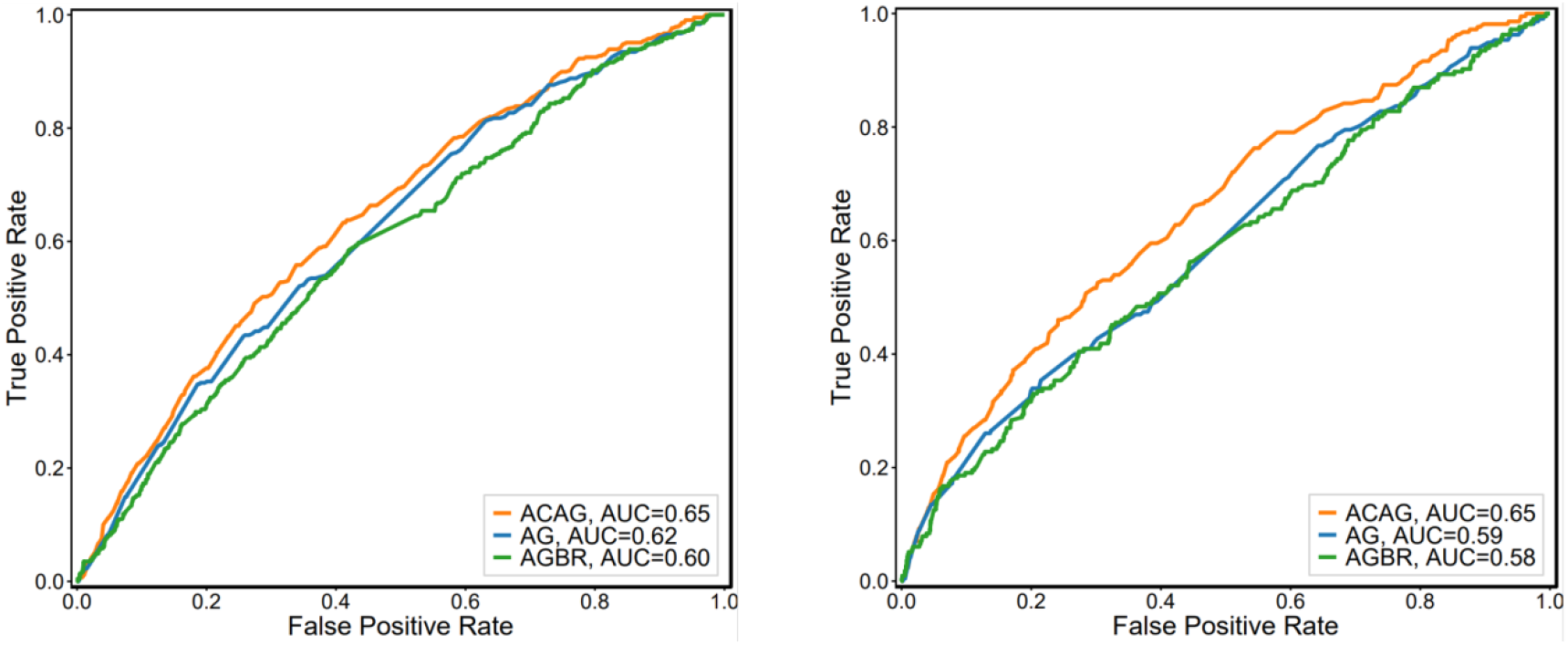
ROC curves for AG-based measurements for predicting DACS (left) and 1-year mortality (right).

## Discussion

In this study, we examined the prognostic value of preoperative ACAG in cardiac surgery patients. Our results showed that higher preoperative ACAG is an independent predictor of DACS and 1-year mortality following cardiac surgery. This finding suggests that preoperative ACAG may improve risk stratification for common post-cardiac surgery complications including delirium and mortality.

Several acknowledged risk factors for DACS were also linked to elevated AG including systemic inflammatory response [29], renal failure [33], and cardiac dysfunction [29]. Therefore, the elevation of AG can serve as a marker of these underlying risk factors, concurrently increasing the patient’s intrinsic susceptibility to developing DACS. Since elevated AG is more likely to be a marker of some underlying cause(s) of DACS, our results suggest that AG is a valuable simple and inexpensive marker for DACS. However, we conjecture that potential interventions derived by elevated AG should focus on addressing the underlying cause of elevated levels of AG not just lowering the AG levels (i.e., via increasing the level of serum bicarbonate).

Recently, Wang et al. [34] demonstrated that postoperative AG, defined as the maximum AG measurement within 24 hours after surgery, is associated with an increased risk of developing DACS. While this finding highlights the prognostic value of AG, its clinical applicability is limited because postoperative measurements do not inform preoperative or intraoperative interventions that could mitigate DACS risk. Our work, relying on preoperative AG offers a more actionable marker, as it reflects the patient’s baseline metabolic state before surgical stress and cardiopulmonary bypass. Elevated preoperative AG may indicate underlying conditions such as lactic acidosis, ketoacidosis, or renal dysfunction, which predispose patients to poor perfusion and systemic inflammation during surgery. Identifying these abnormalities early enables targeted optimization such as correcting metabolic derangements, adjusting fluid strategies, and tailoring perfusion management before irreversible injury occurs. Therefore, preoperative AG is not only predictive but could also provide an opportunity for proactive risk stratification and intervention to reduce the incidence of DACS.

Traditional AG, defined as the difference between unmeasured anions and unmeasured cations, typically using the formula 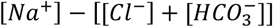. However, serum albumin accounts for the majority of unmeasured anions in the body under normal conditions. Therefore, hypoalbuminemia, which is a common condition among ICU and cardiac surgery patients, yields underestimated AG values that can mask the presence of metabolic acidosis. Following several research studies [23, 28, 29, 35-37], we incorporated ACAG as a more reliable quantifier of the true acid-based status and metabolic derangements. Our results demonstrated that ACAG has a stronger association with DACS and 1-year mortality compared to traditional AG measurement, acknowledging the viability of correcting preoperative AG before using as a prognostic marker for post CABG complications.

Recently, AGBR 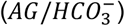 has been explored as a potential marker of metabolic acidosis severity in ICU patients [31]. This ratio integrates both the magnitude of unmeasured anions and the degree of bicarbonate depletion, providing a comprehensive representation of the metabolic acidosis burden and buffering capacity. Our results demonstrated that ACAG provides superior predictive value compared with AGBR. This may be justified, in part, by the inability of AGBR to account for hypoalbuminemia as well as by its sensitivity to variability of 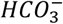 levels due to clinical interventions.

A major strength of this study is that, to the best of our knowledge, it is the first to demonstrate significant association between preoperative ACAG and postoperative delirium in cardiac surgery patients. However, the study has several limitations. First, the data were collected from a single healthcare center. Thus, the reported findings might not generalize to other healthcare systems or other patient populations. Second, the current study is observational in nature. Therefore, despite adjusting for known confounders, the analysis might be biased by some unknown confounders. Despite these limitations, our findings open up opportunities for investigating the use of preoperative ACAG as a valuable, simple, and easily available biomarker of several common post cardiac surgery complications including postoperative delirium and mortality.

## Conclusion

In a retrospective cohort of 4,482 adult patients undergoing cardiac surgery, we found that an elevated preoperative ACAG was significantly associated with increased risk of postoperative delirium and 1-year mortality. Preoperative ACAG is a promising accessible and cost-efficient biomarker that may improve risk stratification for cardiac surgery patients.

## Data Availability

The data from our study has not been deposited into a publicly available repository due to the confidential and sensitive nature of the data including protected heath information.

## Ethics statement

This study was approved by the Institutional Review Board at Geisinger Medical Center, with the approval number: 2020–0101. The need for informed consent was waived by the IRB due to the retrospective nature of the study. The study was conducted in accordance with the Declaration of Helsinki.

